# Somatic development of Wilms tumour via normal kidneys in predisposed children

**DOI:** 10.1101/2024.08.05.24310618

**Authors:** Taryn D. Treger, Jenny Wegert, Anna Wenger, Tim H. H. Coorens, Reem Al-Saadi, Paul Kemps, Jonathan Kennedy, Conor Parks, Nathaniel D. Anderson, Angus Hodder, Aleksandra Letunovska, Hyunchul Jung, Toochi Ogbonnah, Mi K. Trinh, Henry Lee-Six, Guillaume Morcrette, J. Marry M van den Heuvel-Eibrink, Jarno Drost, Ruben van Boxtel, Eline J.M. Bertrums, Bianca F. Goemans, Evangelia Antoniou, Dirk Reinhardt, Heike Streitenberger, Barbara Ziegler, Jack Bartram, J. Ciaran Hutchinson, Gordan Vujanic, Christian Vokuhl, Tanzina Chowdhury, Rhoikos Furtwängler, Norbert Graf, Kathy Pritchard-Jones, Manfred Gessler, Sam Behjati

**Author notes:** These authors contributed equally (Taryn D. Treger, Jenny Wegert, Anna Wenger, Tim H. H. Coorens). Co-corresponding and co-directing authors Contact addresses.

## Abstract

Ten percent of children with cancer harbour a predisposition mutation. In children with the kidney cancer, Wilms tumour, the prevalence is as high as 30%. Certain predispositions are associated with defined histological and clinical features, suggesting differences in tumour genetic development. To investigate this, we assembled a cohort of 137 children with Wilms tumour, of whom 71 had a pathogenic germline or mosaic predisposition. We examined 237 neoplasms (including two secondary leukaemias), utilising whole genome sequencing, RNA sequencing and genome wide methylation, validating our findings in an independent cohort. Tumour development differed between predisposed and sporadic cases, and amongst predisposed children according to specific mutations and their developmental timing. Differences pervaded the repertoire of driver events, including high risk mutations, the clonal architecture of normal kidneys, and the relatedness of neoplasms from the same individual. Predisposition constrains the development of Wilms tumour, suggesting that a variant specific approach to the management of these children merits consideration.

**STATEMENT OF SIGNIFICANCE:** Tumours that arise in children with a cancer predisposition may, or may not, develop through the same mutational pathways as sporadic tumours. We examined this question in the childhood kidney cancer, Wilms tumour. We found that some predispositions strongly constrain the genetic development of tumours, which may have clinical implications.

## INTRODUCTION

Around ten percent of children with cancer are thought to be predisposed to neoplasia^1^. In some entities, such as the kidney cancer Wilms tumour, the proportion has been reported to be as high as 30%^2^. Many genes that have been identified to contribute to Wilms tumour predisposition govern kidney development, with the classical example being *WT1*^3,4^.

Variants may occur in the germline or later in embryogenesis (post-zygotic or mosaic) and be confined to one or more developmental lineages. Traditionally, children with Wilms tumour are screened for predisposing variants if specific clinical features are present, such as young age at diagnosis, bilateral tumours or related congenital anomalies^5^. More recently, predisposition variants have also been identified in children without suspicious features through (unbiased) clinical cancer sequencing programmes^6–8^. Of note, such efforts have also documented cases of Wilms tumour with pathogenic germline mutations in genes predisposing to adult epithelial cancers, such as *CHEK2* or *BRCA2*. Whether or not monoallelic pathogenic germline mutations in these genes are chance findings, or causative in Wilms tumour, remains unclear.

The clinical management of children with a known predisposition differs from that of children with sporadic tumours. According to current protocols of the Children’s Oncology Group (COG) Renal Tumour Committee^9^ and the International Society of Paediatric Oncology Renal Tumour Study Group (SIOP-RTSG)^5^, the aim of treatment is to preserve as much normal kidney tissue as possible, whilst achieving tumour control, given the risk of contralateral tumours and hence, complete kidney loss. Strategies to spare normal kidney tissue include intensifying neoadjuvant (cytotoxic) chemotherapy if needed, nephron sparing rather than total nephrectomy where feasible, and extended courses of post-operative chemotherapy, along with close surveillance for recurrence. Some Wilms tumour predispositions are associated with specific tumour phenotypes. For example, pathogenic germline mutations in *WT1* often lead to tumours exhibiting stromal histology^10^, which predicts favourable oncological outcomes in localised disease. As histology is strongly predictive of disease aggression, it is conceivable that distinctive patterns of somatic mutation link predispositions to specific tumour features, including drivers, high risk features and development of secondary malignancies.

Accordingly, we investigated the genetic development of Wilms tumour from normal kidney tissue in predisposed children, and compared this to sporadic Wilms tumours. We deployed whole genome sequencing, whole transcriptome sequencing and DNA methylation assays across a total of 251 normal tissues and 237 neoplasms derived from 137 children, and validated our findings in a further cohort of 21 children.

## RESULTS

### Overview of discovery cohort

We screened DNA from normal tissues (blood, kidney) of ∼250 children with Wilms tumour to identify individuals with a germline or mosaic cancer predisposition. We obtained tissues from archives assembled in the context of a prospective observational study of childhood renal tumours (IMPORT and UMBRELLA), the SIOP-2001 interventional clinical trial, and institutional archives (**Supplementary Table 1**). We enriched this cohort for children with known predispositions or those at higher risk of predisposition (e.g., children with bilateral tumours or tumours of infants, aged less than 12 months). We subjected blood and kidney DNA to whole genome sequencing (mean coverage 79X) and to methylation arrays, searching for pathogenic germline or mosaic DNA changes in established Wilms tumour predisposition genes (e.g., *WT1*), other cancer predisposition genes (e.g., *TP53*) and cancer genes generally (as per COSMIC (**Supplementary Table 2**))^11–16^. In screening these genes for DNA changes, we considered variation in DNA methylation, substitutions, small indels, copy number changes, as well as rearrangements (including copy number neutral events). With this approach we were able to identify 71 children with a predisposition, which we compared to a cohort of 66 children with Wilms tumour but no identifiable predisposing DNA changes in blood or normal kidney (**Figure 1A; Supplementary Table 1**). The control group was broadly representative of children with Wilms tumour in terms of the two most important independent prognostic factors, tumour histology and stage. The median age was similar across the two groups (39 months for cases; 36 months for controls) (**Supplementary Table 1**). We retrieved all neoplasms (total n = 237) of cases and controls: biopsy (n=1); primary tumour samples (n=219); local or contralateral recurrences (n=6); metastases (n=8); and in two cases, subsequent leukaemias (one lymphoblastic and one myeloid leukaemia) (**Figure 1A; Supplementary Table 1**). We performed whole genome sequencing (n=488 samples; mean coverage 53X), RNA sequencing (n=278/488 samples; 45 million mean reads per sample) and DNA methylation arrays (n=456/488 samples) of neoplasms and normal kidney tissues.

**Figure 1.**
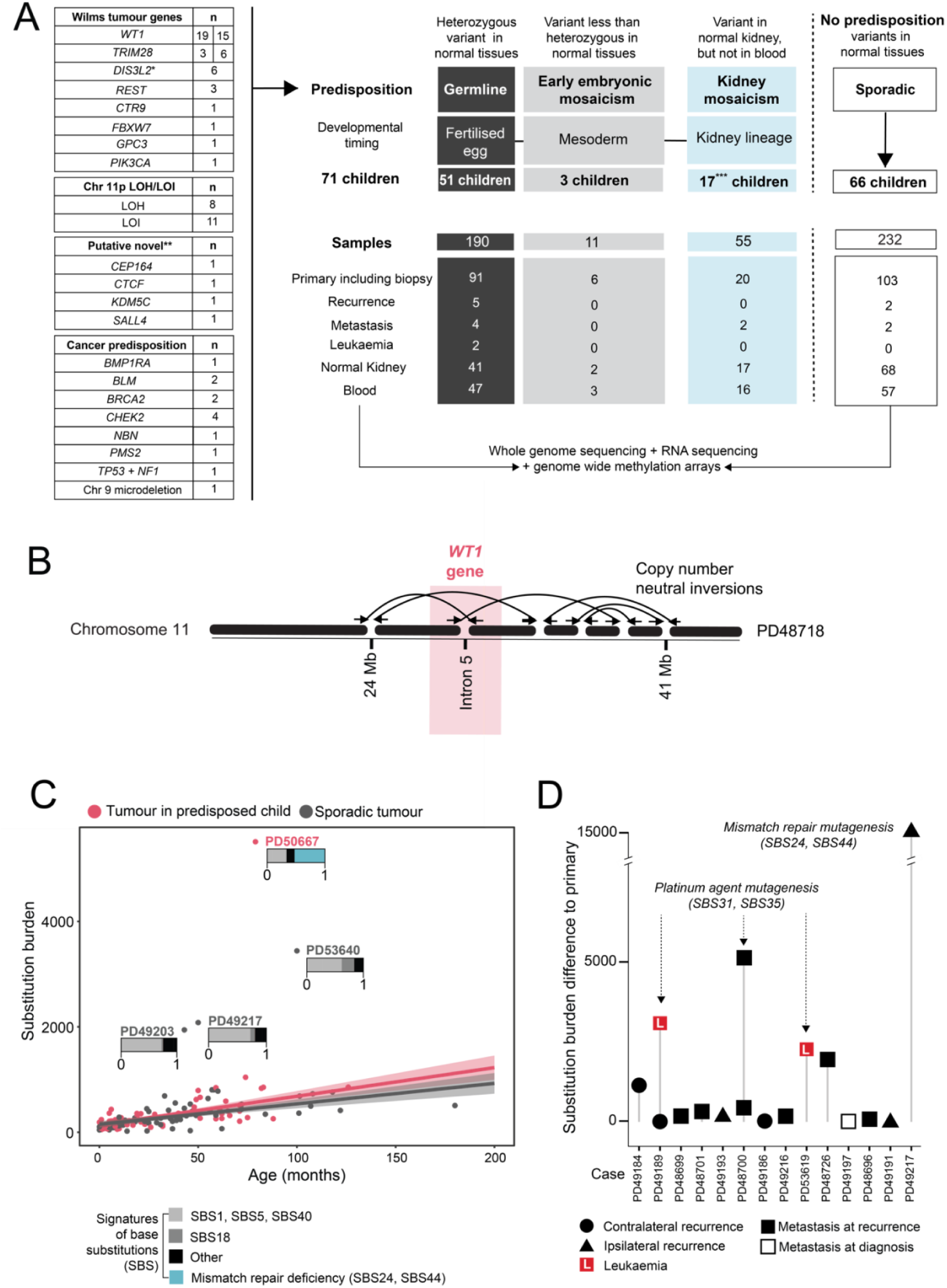
Overview of discovery cohort including substitution burden and signatures. **(A)** Numbers of samples and sample type for each individual. *Monoallelic variant in *DIS3L2,* one child was only registered at recurrence. **Evidence underlying putative novel predispositions is shown in **Supplementary Figure 1 and Supplementary Table 3**. *WT1* and *TRIM28* groups are split into numbers in discovery and validation cohorts. ***1 child did not have blood available for methylation analysis, however phenotypically did not have an overgrowth syndrome. Recurrences included ipsilateral (n=3), contralateral (n=3), and only registered at recurrence (n=1). (**B)** A cryptic complex germline rearrangement overlying *WT1* in an infant (<12 months) with bilateral Wilms tumours. **(C)** Substitution burden in primary tumours from children with or without a predisposition, mutational signatures (stacked bar plots) are shown for outliers. Mutational signatures for entire discovery cohort are shown in **Supplementary Figure 2**. **(D)** Difference between substitution burden for secondary events in relation to matched primary.

### Spectrum of predisposition variants

Genes harbouring predisposition variants fell into one of three categories: 1) established Wilms tumour predisposition changes, including chromosome 11p loss of heterozygosity (LOH)/imprinting (LOI) (n=54 children); 2) cancer predisposition genes that are not usually associated with Wilms tumour (n=13; including one child with two pathogenic germline mutations, in *TP53* (p.R267W) and in *NF1* (p.W784R); 3) novel putative Wilms tumour predisposition genes (n=4 children) (**Figure 1A**). The latter included genes implicated in, but not established as, Wilms predisposition genes. For example, in one child there was a germline deletion of chromosome 16q which encompasses the recessive cancer and putative Wilms predisposition gene, *CTCF*. In all eight samples from the child’s tumour we found a likely disruptive somatic *CTCF* variant of the remaining allele and detected associated hypermethylation of *H19* (thought to be the Wilms tumour promoting effect of *CTCF* loss). Each of these four cases, along with supporting evidence, is shown in **Supplementary Figure 1 and Supplementary Table 3**. Predisposition changes were either germline, acquired post-zygotically in early embryogenesis or were confined to normal kidney tissue, suggesting they arose after kidney parenchymal lineages segregated within the mesoderm from blood lineages (**Figure 1A**). Variants encompassed methylation changes, substitutions, indels, simple and more complex rearrangements. An instructive example was an infant with a suspected predisposition because of bilateral tumours and cryptorchidism, in whom standard of care assays (including germline copy number arrays) had failed to identify a causative predisposition variant. Our approach of looking for structural variants not only through copy number changes, but also through rearrangement calls, identified a series of copy number neutral inversions that disrupted intron 5 of *WT1*, with a truncated transcript, in this child (**Figure 1B**).

### Burden and patterns of somatic variants

We investigated whether differences in the burden and/or patterns of variants may exist that would explain specific features associated with predispositions. We found that the burden of base substitutions correlated with age and did not differ between predisposition cases and sporadic controls (**Figure 1C, 1D**). Substitutions were largely underpinned by mutational processes commonly seen in Wilms tumour (represented in signatures 1, 5, and 18; **Supplementary Figure 2**)^17^. In one primary tumour (PD50667), a disproportionately high mutation burden could be explained by a specific genetic lesion in one case (somatic promoter hypermethylation of *MLH1*). In three other cases (PD49203, PD49217, PD53640, **Figure 1C**), the source of excessive mutations remained cryptic. In recurrences and secondary leukaemia, prior exposure to platinum-based chemotherapy or, in one case acquired mismatch repair deficiency, explained elevated mutation burdens (**Figure 1D**).

Similar to the picture seen in substitutions, we did not find differences in the burden or patterns of indels or rearrangements (as measured by breakpoint burden) (**Supplementary Figure 3**), nor did cases and controls segregate by patterns of methylation (**Supplementary Figure 4**). We did, however, find that the global pattern of methylation strikingly segregated one group of tumours, almost exclusively composed of cases harbouring *WT1* (germline or somatic) and co-mutation of Wnt signalling genes (**Supplementary Figure 4**). As the burden and pattern of somatic variants did not distinguish sporadic from predisposed tumours, any observed differences in subsequent analyses can therefore not be ascribed to a difference in mutation burdens.

### Patterns of somatic driver events

In examining the landscape of driver events, we considered substitutions, indels, copy number changes, rearrangements (both from DNA and RNA sequences), and methylation changes. Focusing on primary tumours (including synchronous contralateral tumours), we identified driver events in cancer genes known to operate in Wilms^12,18^. In addition, we found likely driver mutations in 19 cancer genes not previously implicated in Wilms tumours (**Figure 2A**). Comparing the overall landscape of driver mutations between predisposition cases and sporadic Wilms tumours did not reveal statistically significant differences (**Figure 2B**). However, in children with predispositions involving the *WT1* gene, we did find distinct configurations of driver events. As expected from previous reports^19^, this group harboured somatic chromosome 11p LOH, which includes the *WT1* locus, and driver mutations in genes related to Wnt signalling (mainly *CTNNB1*^20^, including chromosome 3p LOH with homozygosity of mutant *CTNNB1* in some cases, as well as *AMER1*^21,22^), with one related canonical *PIK3CA* mutation^23^. Unexpectedly, there was a striking absence of additional driver mutations (including variants linked to high risk disease) in the *WT1* predisposition group. This observation held true when we assessed multiple biopsies of tumours from the same child, for example PD50643, where we studied six ipsilateral and one contralateral sample, collectively sequenced to 354X. By contrast, in three out of seven sporadic tumours with biallelic somatic *WT1* mutations, we observed a more diverse driver landscape, including subclonal diversification (coloured orange in **Figure 2A**) and one case with a likely high risk variant (chromosome 1q gain, PD50662). Similarly, in another eight predisposed children with mosaic 11p LOH, in whom there was deletion of one copy of *WT1* and duplication of the other copy, driver events emerged in different biological pathways and again, included variants associated with high risk disease.

**Figure 2.**
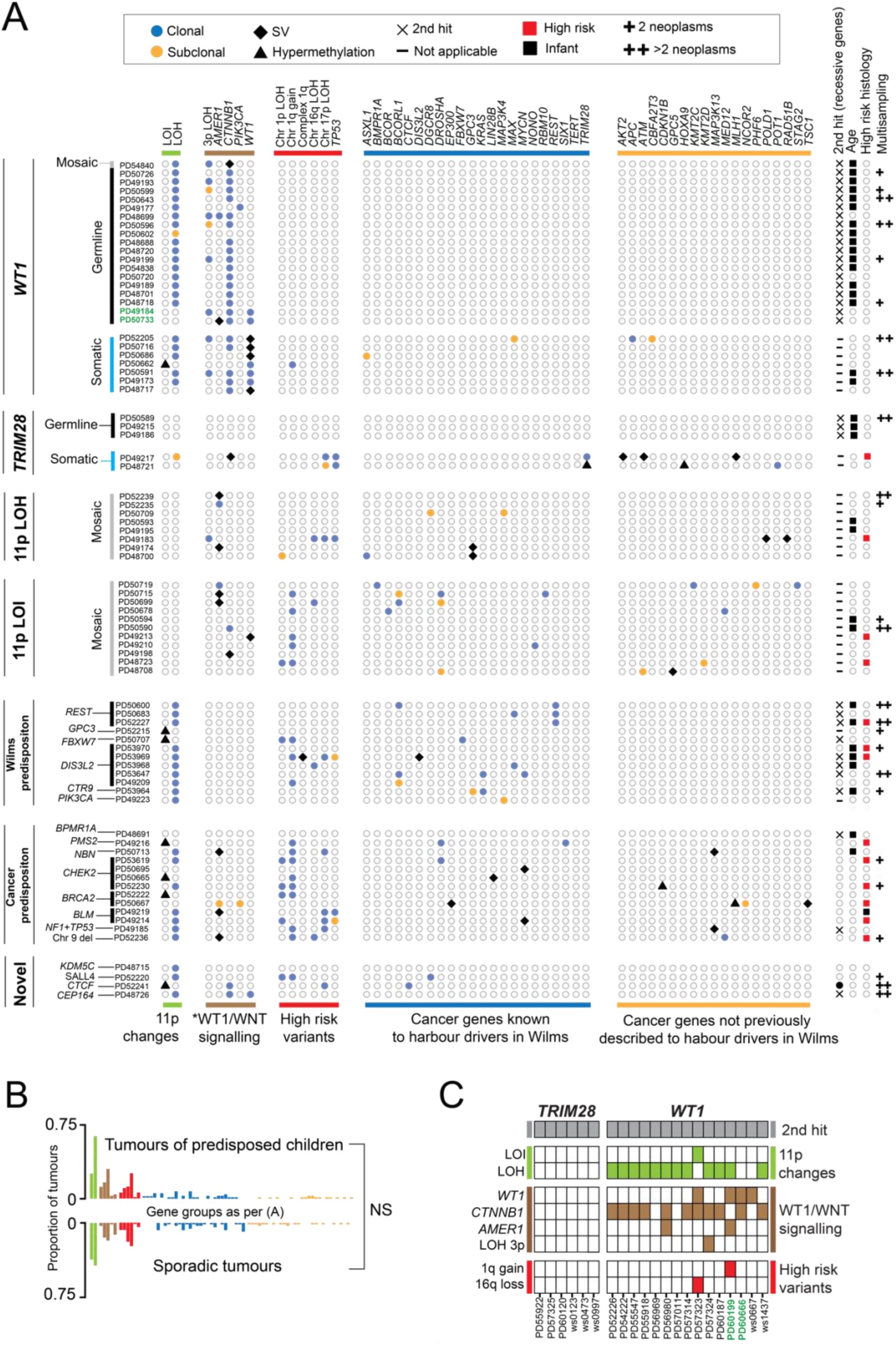
Somatic driver events in predisposition group, sporadic tumours and validation cohort. **(A)** Driver events in primary tumours. Note the confined repertoire of driver events in children with germline *WT1* and germline *TRIM28* events. Multi-sampling and high risk histology are included to highlight that these are not confounding factors. Green highlights children with WAGR syndrome and deletion of chromosome 11p including the *WT1* locus. \**PIK3CA* is included in the Wnt signalling pathway, owing to the crosstalk between Wnt signalling and the PI3K/Akt pathway, converging on *CTNNB1*. Loss of heterozygosity of chromosome 3p, with retention of variant *CTNNB1*, is included as a recurrent event in the group of *WT1*-predisposed tumours. **(B)** Driver events in tumours from children with a predisposition versus children in whom no predisposition could be identified. No significant (NS) difference was found after multiple hypothesis correction testing. **(C)** Driver events in a validation cohort of children with germline *WT1* and germline *TRIM28* recapitulates findings in the discovery cohort. Green highlights children with WAGR syndrome and deletion of chromosome 11p including the *WT1* locus.

The same pattern, albeit derived from a small number of cases, was seen in *TRIM28* driven tumours. In three children with pathogenic germline mutations in the recessive Wilms predisposition gene, *TRIM28*^14^, tumours lost the second copy but did not acquire additional driver events. By contrast, in two non-predisposed children with biallelic *TRIM28* driven tumours, a plethora of driver events emerged somatically, including high risk disease mutations.

A second pattern that emerged was the absence of second “hits” (i.e. driver event) in recessive predisposition genes with an unclear link to Wilms tumour (*NBN*, *PMS2*, *BLM*, *BRCA2*, *CHEK2*), consistent with the aforementioned lack of somatic mutational imprints of the underlying predisposition.

### Validation of preordained driver landscape

To validate the seemingly preordained driver landscape of tumours in *WT1* and *TRIM28* predisposition, we searched for additional cases amongst children with bilateral tumours and/or a family history of Wilms. We thereby identified 15 children with a *WT1* and 6 children with a *TRIM28* germline mutation, interrogating their tumours by whole genome or exome sequencing, methylation arrays and RNA sequencing (**Figure 2C**). Consistent with our previous findings, each tumour exhibited a second mutation in the underlying predisposition gene and in cases of *WT1* predisposition, driver variants in Wnt signalling genes. Otherwise, there was a paucity of additional driver variants except for in two cases. In one child with WAGR syndrome, caused by deletion of not only *WT1* but also neighbouring genes including *PAX6*, the Wilms tumour harboured gain of chromosome 1. In another child, who presented at an unusually old age (50.1 months vs. 15-19 months for other children with *WT1* germline mutations), we found somatic loss of chromosome 16q. Overall, the absence of additional driver variants in tumours arising on a background of *WT1* or *TRIM28* predisposition was significant, even when solely considering high risk variants, whether compared to other predispositions (p=0.0001) or to control tumours (p<0.05, Fisher’s exact test).

### Effects of predisposition on the clonal architecture of normal kidney tissue

We have previously described clonal expansions in normal kidney tissue, which were direct precursors of Wilms tumours^24^. The expansions were measurable as kidney-specific mutations (relative to blood), associated in some cases with the Wilms predisposition variant, hypermethylation of *H19*. To investigate whether predisposition generally distorts the clonal architecture of normal kidney tissue, we searched for variants in normal kidney (relative to blood), subdividing these into mutations that precede tumours and those that are not shared with tumour (**Figure 3A)**. This analysis showed that normal kidney clonal expansions occur not only in mosaic hypermethylation of *H19* but also in kidney-specific mosaic chromosome 11p LOH in children without any reported phenotype of overgrowth syndrome. The occurrence of clonal expansions varied amongst children with other predispositions and in sporadic tumours. Moreover, the degree to which these expansions perturbed normal kidney tissue, as inferred from the largest variant allele frequency of the clone and the number of mutations defining each clone, was statistically greater in tumours arising on a background of mosaic 11p changes (**Figure 3B, 3C**). Overall, these findings indicate that, whilst distortions of the clonal architecture of normal kidneys are not ubiquitous, they do occur across the spectrum of Wilms tumours. The greatest impact is seen in cases of 11pLOH and *H19* hypermethylation.

**Figure 3.**
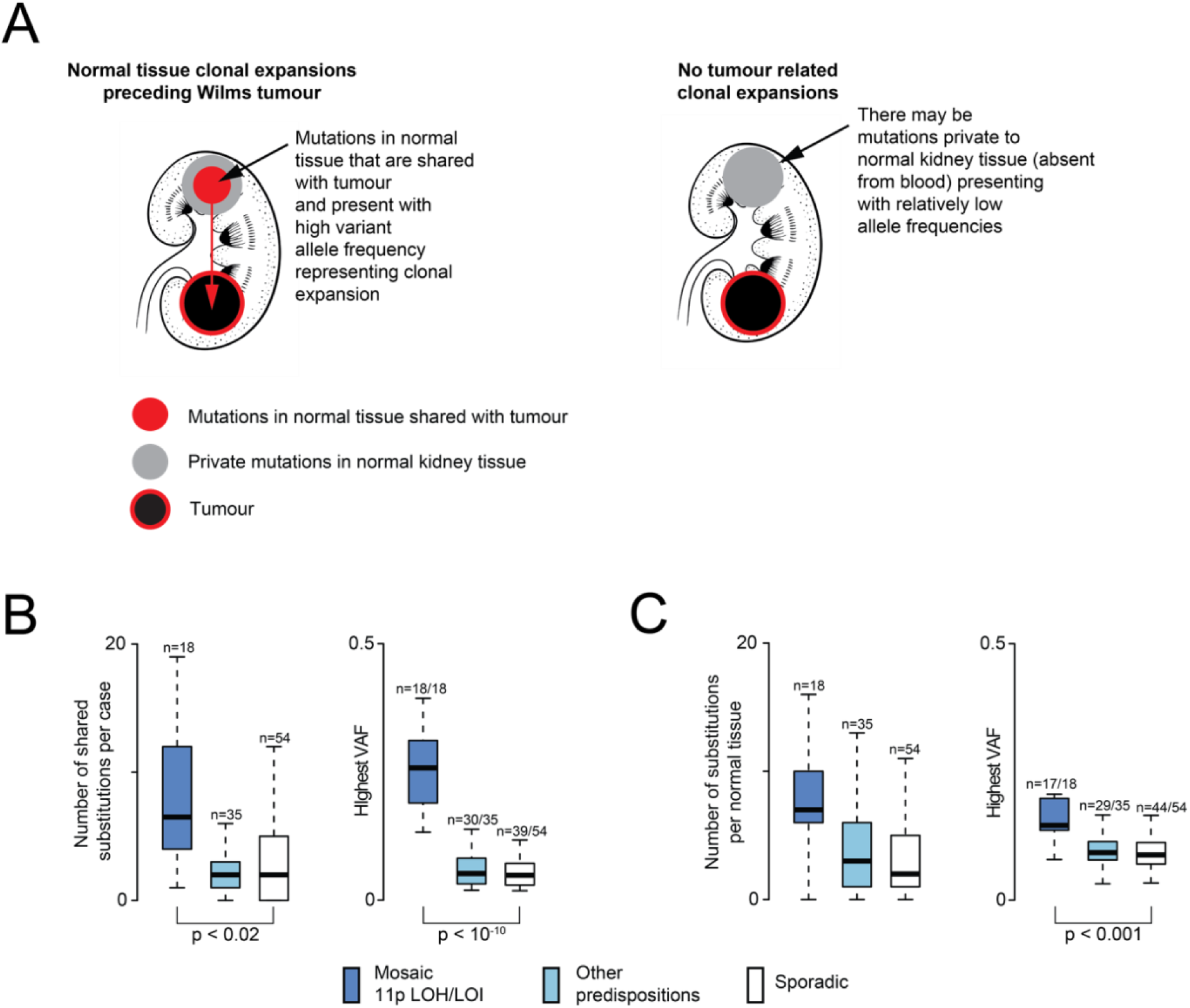
Clonal expansions in normal kidney tissue. **(A)** Schematic to illustrate the concept of clonal expansions in normal kidneys. **(B)** Boxplot of number of substitutions shared between normal kidney tissue and tumour for each case and of their variant allele frequencies (VAF). **(C)** Boxplot of number of private (i.e. not shared with tumour) substitutions in normal kidney tissue sample for each case and of their variant allele frequencies (VAF).

### Phylogenetic relation of multiple neoplasms of the same individual

In 20 children we were able to examine more than one neoplasm including bilateral tumours, recurrences, metastases, and leukaemias. We examined not only cancer evolution (i.e. the phylogenetic relationship of somatic mutations across different neoplastic samples) but also embryonic development by considering mosaic mutations shared between tumours and normal tissues (**Figure 4A**, **Figure 4B**). Using this approach, we reconstructed the development of each neoplasm (**Supplementary Figure 5, Supplementary Figure 6**). An overview of the cancer evolution of the investigated neoplasms is shown in **Figure 4C**. Consistent with previous reports^19^, we found that the development of bilateral tumours may be independent, whereas multiple lesions from the same tumour bulk shared a common origin. Instructive examples were two children in whom we sampled bilateral tumours as well as multiple biopsies from the one side (PD50643, *WT1* predisposition; PD52239, chromosome 11p LOH). Left and right tumours did not share any (non-embryonic) somatic variants whereas tumour biopsies taken from the same side developed from a common tumour trunk. We were able to show that this pattern of independence extended to recurrences. Ipsilateral recurrences, but not contralateral recurrences, shared a mutational trunk with the primary tumour. This pattern of independence even applied to metastatic lesions. In three predisposed children, metastases emerged via an independent phylogenetic route. For example, in a child with a *WT1* germline predisposition and bilateral tumours at diagnosis (PD48701), a metastatic lung recurrence shared no somatic mutations with the primary tumour, and had segregated early in development, as evidenced by mutually exclusive mosaic mutations (**Figure 4D, 4E**). Whilst the metastasis likely arose from the unsampled contralateral primary, without examining mosaic mutations, one might have concluded that the metastatic recurrence derived from the sampled primary, given they superficially share 11p LOH (with different breakpoints) and the same *CTNNB1* hotspot mutation (**Figure 4E, 4F**). Finally, we examined two instances of secondary leukaemia after Wilms tumour (**Figure 4D, 4E, 4F**). A child (PD49189) with a *WT1* germline mutation and bilateral asynchronous Wilms tumours developed acute myeloid leukaemia. Leukaemic clones had no phylogenetic relation to either Wilms tumour. They exhibited platinum agent mutagenesis (from Wilms treatment), leukaemogenic drivers and somatic hits in *WT1* (truncating variant and subclonal 11p LOH), supporting a causative role of the germline *WT1* mutation in the AML of this child. The second case was a child (PD53619) with a pathogenic germline *CHEK2* mutation and unilateral Wilms. Unusually for a secondary leukaemia, it was lymphoblastic rather than myeloid. Again, the leukaemia bore mutational imprints of platinum agent exposure (from Wilms treatment) and leukaemogenic driver variants, with no phylogenetic relation to the tumour or its embryonic root. Neither the leukaemia nor the Wilms tumour of this child harboured a second hit in the recessive cancer gene, *CHEK2*.

**Figure 4.**
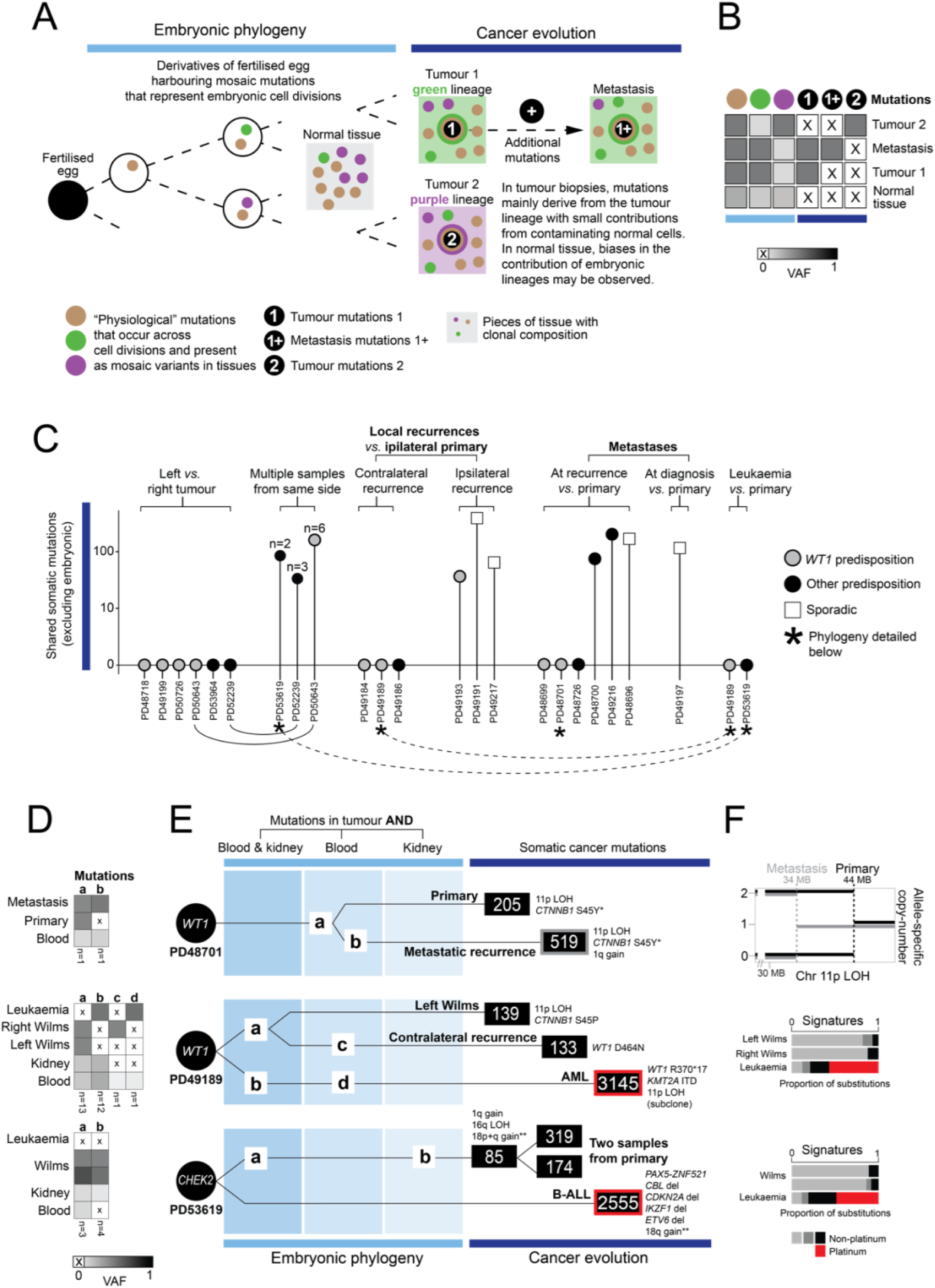
Phylogenetic relationship of blood, normal kidney and multiple tumours in predisposed children. **(A)** Schematic to illustrate phylogenetic reconstruction. **(B)** Variant allele frequency (VAF) heatmap of embryonic and tumour somatic mutations, across normal and tumour biopsies, to illustrate the VAF inflation of embryonic mutations in tumours via clonal expansions. **(C)** The sharedness of tumour somatic mutations between primary and secondary events in children with *WT1* predisposition, other predisposition and sporadic cases. **(D)** VAF heatmap of embryonic mutations across all samples from an individual, underlying phylogeny reconstruction shown in (E). Letters correspond to letter in (E). The number at the bottom of each column refers to the number of mutations represented by each letter. **(E)** Example phylogenies. Circle represents zygote with predisposition mutations. Patient ID underneath. Lines (not scaled to mutation burden) represent phylogenetic relations. Number in squares are the tumour substitution burden. Variants listed are the driver events of each neoplasm. *Same hotspot mutation in *CTNNB1* that has evolved in parallel (explained in text). **18q gain is shared between the three tumour samples but with different breakpoints in each sample. It is not a common copy number change in either Wilms tumour or ALL **(F)** Noteworthy genomic features. Top: Different breakpoints in 11p LOH in both tumours. Middle and bottom – signatures of substitutions highlighting mutations due to platinum chemotherapy agents (red) in secondary leukaemias.

## DISCUSSION

We examined the somatic evolution of Wilms tumour via normal kidney tissue in predisposed children. Our principal observation was that some predispositions seem to restrict and dictate the somatic evolution of tumours. Our findings therefore provide a rationale for exploring a variant-specific approach to the clinical management of predisposed children with Wilms tumour.

We found a restrained driver landscape in children with pathogenic *WT1* mutations which was, in particular, devoid of variants representing either high risk Wilms tumour (*TP53*, 17p loss) or those currently used by COG to intensify treatment (combined LOH of 1p and 16q, 1q gain). This observation provides an explanation for the established association of *WT1* germline mutations with non high risk histology^25^. For example, diffuse anaplasia, an unambiguous histological feature of aggressive Wilms tumour, has been firmly linked to *TP53* mutation^26,27^. The complete absence then of *TP53* mutation in any of the 72 tumours (from 34 children with a predisposing *WT1* event) provide a genetic explanation for the exceptional rarity of *TP53*-associated high risk histology (i.e. diffuse anaplasia) in primary tumours of children with germline *WT1* mutations. By contrast, in tumours with somatic *WT1* mutations and in chromosome 11p LOH, the driver landscape was less restrained.

Similarly, our finding of the absence of additional driver variants, in particular of high risk mutations, in children with pathogenic *TRIM28* germline mutations explains why diffuse anaplasia in this group is exceedingly rare^28^. Overall, these findings indicate that the developmental timing and nature of the predisposing mutation may predetermine a somatic evolutionary trajectory.

Our finding of predisposition-specific somatic patterns extended to the clonal architecture of normal tissue. In children with chromosome 11p changes (LOH or hypermethylation), we found large clonal expansions in normal kidney tissues that preceded tumour development, whereas in other predispositions and in sporadic tumours such expansions were either much less pronounced or absent. Whilst this does not exclude the presence of large clonal expansions in regions of the kidney we did not sample, our inability to detect these suggests that Wilms germline predispositions do not necessarily perturb the clonal architecture of normal kidneys. Mosaic predispositions, by contrast, commonly and more profoundly altered the clonal architecture of kidneys. This would suggest that mosaic chromosome 11p changes do not merely represent a polyclonal field of cancer prone cells. Instead, they represent the first (i.e. predisposition) and second (i.e. clonal outgrowth) steps of tumour evolution. By contrast, germline predispositions “only” generate cancer prone cells without a clonal expansion in normal tissues. Finally, our detailed phylogenetic analyses, which included early embryonic development, illustrate varied instances of independent tumour generation in predisposed children.

We included children with pathogenic germline mutations that have unknown links to Wilms tumour formation in the context of germline heterozygosity: *BRCA2, CHEK2, BLM* and *PMS2.* Pathogenic germline mutations in these genes have been repeatedly documented in children with a variety of cancer types^29^, but the degree of pathogenicity remains unclear. We did not find evidence that haploinsufficiency of these genes influenced tumour genomes, which does not exclude the possibility that these mutations increased cancer risk, either in isolation or in collaboration with other variants. To answer the question of pathogenicity, we would suggest that population based studies are required to establish whether haploinsufficiency of these genes is significantly enriched in children with Wilms tumour.

In managing predisposed children with Wilms, we have to balance the imperative for tumour eradication with the need to preserve renal function. Measures to achieve this include partial, rather than radical, nephrectomies, aiming for maximum nephron-sparing surgery whilst minimising the risk of positive resection margins. The overall clinical inference from our findings is that children with a predisposition may potentially benefit from a variant specific overall management plan. We may consider, for instance, extending neoadjuvant treatment in children with pathogenic *WT1* or *TRIM28* germline mutations, given the seemingly low incidence of high risk disease mutations at diagnosis. In the case of *WT1* driven tumours, which are often stromal in histology and therefore difficult to shrink with cytotoxic agents^30^, this may require the development of novel agents. Furthermore, we could explore whether distortions of the clonal architecture of normal kidneys correlate with recurrence risk, which could then guide the need for, and duration of, both surveillance and maintenance chemotherapy for secondary prevention. Finally, it may be possible that the intensity of relapse therapy considers the underlying phylogenetic configuration, i.e. whether a relapse evolved independently or is a direct descendant of the original tumour. Pursuing these hypotheses will require international efforts to assemble statistically meaningful patient cohorts, coupled with genomic studies of tumour and surrounding normal tissues.

*Bonafide* childhood cancer predisposing mutations are relatively common in paediatric oncological practice. Our findings in Wilms tumour indicate that predispositions may forge the genetic development of neoplasms. We would therefore encourage similar investigations into other cancer types to elucidate the nuances and common principles that underpin tumour evolution in predisposed children.

∼

The authors declare no potential conflicts of interest.

## Acknowledgements

This study is part of The Little Princess Trust Knowledge Bank of Wilms Tumour funded by The Little Princess Trust. Additional funding was provided by the Wellcome Trust (personal fellowship to S.B., institutional grant to the Wellcome Sanger Institute; references 220540/Z/20/A and 223135/Z/21/Z), the Wenner-Gren Foundations (personal fellowship to A.W.) and the Pessoa de Araujo family (personal fellowship to A.H.). This research was supported by the NIHR GOSH Biomedical Research Centre and NIHR Cambridge Biomedical Research Centre (NIHR203312). Within Germany the SIOP 2001 study and trial was supported by Deutsche Krebshilfe (grant 50-2709-Gr2). Work in the lab of M.G. was supported by the DFG (Ge539) and the Wilhelm-Sander-Stiftung. The views expressed are those of the authors and not necessarily those of the NHS, the NIHR or the Department of Health. We are indebted to the children and families who participated in this study.

## Author Contributions

S.B., T.D.T. and T.H.H.C. designed the experiment. T.D.T. performed overall data analysis aided by J.W. (extension cohort), A.W. (phylogenetic analyses, methylation clustering), and T.H.H.C. (signature analyses; analysis of normal tissue clones). P.K., N.D.A., A.H., H.J. M.T. further contributed to data analysis. The following authors prepared samples and / or provided clinical data and expertise and / or prepared experiments: J.W., R.A.S., J.K., C.P., T.O., A.L., J.D., R.v.N., E.B., B.G., M.vdHE., E.A., D.R., B.Z., H.S., J.B., R.F., N.G., T.C. Pathology expertise was provided by H.L.S., G.M., C.H., G.V., and C.V. K.P.J., M.G., and S.B. co-directed the study. S.B. and T.D.T. wrote the manuscript.

## Data availability

De-identified patient level information and all variants calls are provided in Supplementary Tables. Raw DNA data generated in this study have been deposited in the European Genome-Phenome Archive (EGA) under accession code EGAS00001004237. Raw RNA data generated in this study have been deposited in the European Genome-Phenome Archive (EGA) under accession code EGAS00001005244.

## MATERIALS AND METHODS

### Patients and Sampling

We obtained samples from these sources: the UK Improving Population Outcomes for Renal Tumours of Childhood (IMPORT) study, now known as UMBRELLA (approved by an NHS research ethics committee, references London Bridge REC 12/LO/0101); the German arm of the SIOP 2001 study (EudraCT number SIOP 2001: <u>2007-004591-39</u>, approved by Ethik-Kommission der Ärztekammer des Saarlandes, approval numbers 136/01 and 248/13); and the Netherlands (approved by Medisch Ethische Toestings Commissie ErasmucMC, approval numbers MEC 202.134/2001/122, MEC-2018-026, MEC-2006-348 and Netherlands Trial Register NL7744 MEC-2016-739). Institutional archive and biobank consent was provided by the institutional biobank committee of the Princes Máxima Centre, Utrecht. PD IDs are de-identified and unknown to anyone.

Patients or their legal guardians had provided consent to research. Patients over 6 months were treated with preoperative chemotherapy regimens according to stage, as per SIOP WT 2001. Patients under 6 months underwent upfront nephrectomy or nephron sparing surgery. Following surgery, patients were risk stratified to subgroups based on histology, and postoperative treatment was further refined according to stage of disease at surgery. Normal kidney and tumour tissues were collected at surgery, and blood throughout treatment. Normal kidney was sampled by pathologists according to the SIOP-RTSG standard biobanking protocols, from a morphologically normal appearing corticomedullary region distant from the tumour.

### Nucleic Acid Extraction

Samples used for DNA/RNA extraction were fresh-frozen stored at −80°C. Tumour and normal kidney DNA/RNA were extracted by standard methods using the Qiagen Allprep kit/QIAmp DNA kit/RNeasy kit. Blood germline DNA was extracted with the QIAamp DNA Blood Midi Kit (Qiagen).

### DNA Sequencing and Alignment

Short insert (500bp) genomic libraries were constructed, flowcells prepared and 150 bp paired-end sequencing clusters generated on the Illumina HiSeq X platform. Tumours were sequenced to an average depth of 53X, normal kidney to 82X and blood to 75X. DNA sequences were aligned to the GRCh37 reference genome by the Burrows-Wheeler algorithm (BWA-MEM)^31^.

### Detection of Variants

Using the extensively validated analysis pipeline of the Wellcome Sanger Institute, all classes of mutations were called: substitutions (CaVEMan), indels (Pindel), copy number variation (ASCAT and Battenberg) and rearrangements (BRASS)^32–35^. The CaVEMan, Pindel and BRASS algorithms were run matched (tumour versus matched normal) and unmatched (all samples against an in silico human reference genome). Mapping artefacts were removed by setting a threshold for the median alignment score of reads supporting a variant (ASMD>=140) and requiring that fewer than half of the reads were clipped (CLPM=0). Pindel variants were filtered out if the quality score was below 300. Rearrangements were validated if meeting the criteria of either an assembly score or minimum number of reads (4 in tumours, 25 in normal samples).

### Germline and somatic filtering

A binomial distribution was fitted to the combined read counts of all normal samples from one individual per substitution or indel, with the depth as the number of trials, and the number of reads supporting the variant as number of successes^24^. Germline and somatic variants were differentiated based on a one-sided exact binomial test. The null hypothesis is that the number of reads supporting the variants, across copy number normal samples, is drawn from a binomial distribution with p=0.5 (p=0.95 for sex chromosomes in males), and the alternative hypothesis drawn from a distribution with p<10^-5^ to minimise false positives. Variants for which the null hypothesis could be rejected were classified as somatic (embryonic mosaic or tumour), otherwise as germline. Additional filtering involved removing substitutions within 10bp of an indel, and variants were also filtered out if they were called in a region of consistently low or high depth across all diploid samples from one individual. All variants detected from samples belonging to the same individual were then recounted twice: once with a threshold of read mapping quality 30 and base quality 25, defining high-quality reads, and once without applying thresholds for mapping and base quality. Variants were included if the fraction of high-quality reads over total reads was greater than 75%.

### Annotation of Somatic Driver Events

Variants in genes considered cancer genes were annotated, as per Tier 1 genes from the Census of Cancer Genes and from a curated list of known Wilms tumour drivers^12,16,18^. Missense mutations and in frame indels were considered drivers if occurring in canonical hot spots of oncogenes. As TERT promoter mutations fail Caveman’s PASS filter due to being localised in area of simple repeats, these mutations were manually rescued. Truncating mutations were considered drivers if predicted to disrupt the footprint of recessive cancer genes. Focal (< 1 MB) homozygous deletions and amplifications (copy number > 4 (diploid) or 8 (tetraploid)) in recessive and dominant cancer genes, respectively, were considered drivers. Sub amplifications in oncogenes were included if accompanied by significantly high RNA expression, as determined by z score. Rearrangements were considered driver events when they generate a known oncogenic gene fusion or when their breakpoints disrupted the gene footprint of recessive genes. Rearrangements affecting regulatory domains of oncogenes were included if RNA expression was elevated, as per z score.

Rearrangements were validated by manual inspection of both WGS and RNA sequencing data on the genome browser JBrowse to exclude further sequencing artefacts^36^. Tumour purity, local copy number state and variant allele fraction (VAF) was used to calculate cancer cell fraction^37^ in order to differentiate between clonal and subclonal substitutions/indels. Clonality for copy number changes was determined from Battenberg.

### Annotation of Germline and Mosaic Predisposition Events

Variants in genes considered predisposition genes were annotated, as per the Census of Cancer Genes, from a curated list of known Wilms tumour predisposition genes or if identified as pathogenic in a large scale whole genome/exome sequencing study of 1120 children with cancer^11–16^. Common variants were discounted if listed on dbSNP with an allele frequency >0.001, unless reported to be pathogenic, or if identified as a recurrent structural variant in gnomAD^38,39^. Substitutions were also removed if predicted to be benign and tolerated by SIFT and PolyPhen-2 respectively^40,41^. Null monoallelic variants in recessive genes were included if previously reported to be dominant negative or haploinsufficient, or if occurring on the X chromosome. Else, monoallelic variants affecting recessive genes were disregarded if there was no second somatic hit, either via a substitution, indel, rearrangement, deletion or hypermethylation of the promoter. All potential variants were cross-referenced with mutations catalogued in the Human Gene Mutation Database Professional (portal.biobase-international.com), ClinVar (https://www.ncbi.nlm.nih.gov/clinvar/) and VarSome (https://varsome.com) as predisposing to Wilms tumour or identified as pathogenic in hereditary cancers.

### Screening for mosaic or germline 11p loss of heterozygosity

To identify mosaic or germline copy number changes, raw ASCAT^34^ counts from blood and normal kidney were subset based on the coordinates of 11p loss of heterozygosity (LOH) within the tumour. The raw ASCAT data was converted to VAF by dividing the counts for each nucleotide at each position by the depth of each read. The alternate nucleotide was defined from homozygous SNPs within the region of LOH in the tumour. For any normal samples with a mean VAF > 0.5 within the 11p LOH region, plots of B-allele frequency were manually inspected.

### Clonal precursors

We assessed normal kidney for the presence of clonal expansions as previously described^24^, by interrogating the normal blood and normal kidney samples of patients for somatic substitutions shared with the Wilms tumour. Putative clonal precursors to Wilms tumours were identified as substitutions present in both the Wilms tumour and normal kidney, but absent from blood, defined as less than two supporting reads. In addition, we looked for substitutions in normal kidney samples that were absent from blood. All variants were manually inspected using the genome browser JBrowse^36^.

### Phylogenies

Variants in regions where all tumour samples were non-diploid were excluded from phylogenetic analyses. We employed a beta-binomial model to separate true somatic mutations from noise in each sample^24^. The locus-specific error rate was calculated using blood samples from 50 children with sporadic Wilms as the reference panel. The resulting p-values were corrected for multiple testing using the Benjamini-Hochberg procedure^42^ and a cut-off was set at q<0.005. Remaining variants shared between at least two samples from an individual were manually inspected using the genome browser JBrowse^36^ to exclude artefacts, before classifying mutations as unique or shared between samples based on VAF and tumour purity. The phylogenetic relationships between tumours and normal tissues were then reconstructed based on their shared embryonic and somatic mutations, as done previously^43–45^.

### Mutational Signature Analysis

We extracted signatures of base substitutions using the hierarchical Dirichlet process (HDP) algorithm, as previously employed^46^. We used the patients as hierarchy and aggregated results of 20 independent Markov Chain Monte Carlo runs, each taking 100 samples from 40,000 iterations with the first 20,000 as burn-in. As before, the resulting HDP signatures were then deconvolved into linear mixtures of COSMIC reference signatures using an expectation maximization algorithm. Signatures that had a deconvolution with a cosine similarity greater than 0.8 were broken down into their constituent reference signatures. If not, HDP signatures were kept without deconvolution.

### Bulk RNA Analysis

RNA libraries were sequenced on the Illumina HiSeq 4000 platform. Reads were aligned using STAR, and mapped to GRCh37, and read counts of genes were obtained using HTSeq^47,48^. Data was processed in R using Edge R, normalised using the TMM method and converted to log-CPM values^49^. Bwcat (version 1.5.2), from the in-house cgpBigWig package, was used to determine raw coverage across genes of interest and then normalised per exon.

### Methylation Analysis

Data on genome-wide methylation status was obtained using the Illumina Infinium MethylationEPIC BeadChip microarray kit. Data was processed and normalised using the Funnorm method in R using Minfi^50^. Comparisons were made using the beta score, which is the ratio of intensities between methylated and unmethylated alleles. Hypermethylation of *H19* was explored using probes in ICR1 (chr11:2018812-2024740), and compared to probes within ICR2 (chr11: 2629558-2721224). Promoter hypermethylation of 318 Census tumour suppressor genes^16^ and Wilms tumour recessive genes^13–15^ was assessed using promoter-associated probes or, if these were not annotated, probes within the 5’UTR, 1^st^ exon or transcription start sites of the genes. Beta values for promoters were compared to log-cpm values for RNA expression, and any sample with both a high beta value (as per z score >=3) and low expression (as per z score <=3) were annotated as having promoter hypermethylation.

For methylation clustering, we included all tumours samples with a tumour cell content above 60%, as estimated by Battenberg^35^. For children with a multi-sampled tumour, we only included the sample with the highest purity. In total, 140 tumour samples from 121 children (67 predisposed and 54 sporadic) were used for methylation clustering. The common methylation probes for all 140 samples were selected, excluding probes in the imprinted gene *KCNQ1DN*. The Euclidean distance between samples was calculated based on the 10000 most variable CpG sites, before unsupervised clustering. A Fisher exact test was performed to test the dependency of a *WT1* alteration in addition to a *CTNNB1*, *AMER1* or *PIK3CA* alteration in tumours belonging in methylation cluster 1 (the leftmost cluster).

## REFERENCES

1. Kratz, C. P. et al. Predisposition to cancer in children and adolescents. Lancet Child Adolesc Health 5, 142–154 (2021).

2. Hol, J. A. et al. Prevalence of (Epi)genetic Predisposing Factors in a 5-Year Unselected National Wilms Tumor Cohort: A Comprehensive Clinical and Genomic Characterization. J. Clin. Oncol. 40, 1892–1902 (2022).

3. Pelletier, J. et al. WT1 mutations contribute to abnormal genital system development and hereditary Wilms’ tumour. Nature 353, 431–434 (1991).

4. Pritchard-Jones, K. et al. The candidate Wilms’ tumour gene is involved in genitourinary development. Nature 346, 194–197 (1990).

5. Vujanić, G. M. et al. The UMBRELLA SIOP–RTSG 2016 Wilms tumour pathology and molecular biology protocol. Nat. Rev. Urol. 15, 693–701 (2018).

6. Bakhuizen, J. J. et al. Assessment of Cancer Predisposition Syndromes in a National Cohort of Children With a Neoplasm. JAMA Netw Open 6, e2254157 (2023).

7. Villani, A. et al. The clinical utility of integrative genomics in childhood cancer extends beyond targetable mutations. Nat Cancer 4, 203–221 (2023).

8. Shukla, N. et al. Feasibility of whole genome and transcriptome profiling in pediatric and young adult cancers. Nat. Commun. 13, 2485 (2022).

9. Chintagumpala, M. M. et al. Outcomes based on histopathologic response to preoperative chemotherapy in children with bilateral Wilms tumor: A prospective study (COG AREN0534). Cancer 128, 2493–2503 (2022).

10. Schumacher, V. et al. Correlation of germ-line mutations and two-hit inactivation of the WT1 gene with Wilms tumors of stromal-predominant histology. Proceedings of the National Academy of Sciences 94, 3972–3977 (1997).

11. Zhang, J. et al. Germline Mutations in Predisposition Genes in Pediatric Cancer. N. Engl. J. Med. 373, 2336–2346 (2015).

12. Treger, T. D., Chowdhury, T., Pritchard-Jones, K. & Behjati, S. The genetic changes of Wilms tumour. Nat. Rev. Nephrol. (2019) doi:10.1038/s41581-019-0112-0.

13. Mahamdallie, S. et al. Identification of new Wilms tumour predisposition genes: an exome sequencing study. The Lancet. Child & Adolescent Health 3, 322 (2019).

14. Halliday, B. J. et al. Germline mutations and somatic inactivation of TRIM28 in Wilms tumour. PLoS Genet. 14, e1007399 (2018).

15. Mahamdallie, S. S. et al. Mutations in the transcriptional repressor REST predispose to Wilms tumor. Nat. Genet. 47, 1471–1474 (2015).

16. Tate, J. G. et al. COSMIC: the Catalogue Of Somatic Mutations In Cancer. Nucleic Acids Res. 47, D941–D947 (2019).

17. Thatikonda, V. et al. Comprehensive analysis of mutational signatures reveals distinct patterns and molecular processes across 27 pediatric cancers. Nature Cancer 2023 4:2 4, 276–289 (2023).

18. Gadd, S. et al. A Children’s Oncology Group and TARGET initiative exploring the genetic landscape of Wilms tumor. Nat. Genet. 49, 1487–1494 (2017).

19. Murphy, A. J. et al. Genetic and epigenetic features of bilateral Wilms tumor predisposition in patients from the Children’s Oncology Group AREN18B5-Q. Nat. Commun. 14, 8006 (2023).

20. Li, C.-M. et al. CTNNB1 mutations and overexpression of Wnt/beta-catenin target genes in WT1-mutant Wilms’ tumors. Am. J. Pathol. 165, 1943–1953 (2004).

21. Ruteshouser, E. C., Robinson, S. M. & Huff, V. Wilms tumor genetics: mutations in WT1, WTX, and CTNNB1 account for only about one-third of tumors. Genes Chromosomes Cancer 47, 461–470 (2008).

22. Wegert, J. et al. WTX inactivation is a frequent, but late event in Wilms tumors without apparent clinical impact. Genes Chromosomes Cancer 48, 1102–1111 (2009).

23. Prossomariti, A., Piazzi, G., Alquati, C. & Ricciardiello, L. Are Wnt/β-Catenin and PI3K/AKT/mTORC1 Distinct Pathways in Colorectal Cancer? Cell Mol Gastroenterol Hepatol 10, 491–506 (2020).

24. Coorens, T. H. H. et al. Embryonal precursors of Wilms tumor. Science 366, 1247 (2019).

25. Falcone, M. P. et al. Long-term kidney function in children with Wilms tumour and constitutional WT1 pathogenic variant. Pediatr. Nephrol. 37, 821–832 (2022).

26. Maschietto, M. et al. TP53 mutational status is a potential marker for risk stratification in Wilms tumour with diffuse anaplasia. PLoS One 9, e109924 (2014).

27. Wegert, J. et al. TP53 alterations in Wilms tumour represent progression events with strong intratumour heterogeneity that are closely linked but not limited to anaplasia. Hip Int. 3, 234– 248 (2017).

28. Wegert, J. et al. TRIM28 inactivation in epithelial nephroblastoma is frequent and often associated with predisposing TRIM28 germline variants. J. Pathol. 262, 10–21 (2024).

29. Capasso, M. et al. Genetic Predisposition to Solid Pediatric Cancers. Front. Oncol. 10, 590033 (2020).

30. Duncan, C. et al. Response of bilateral wilms tumor to chemotherapy suggests histologic subtype and guides treatment. J. Natl. Cancer Inst. (2024) doi:10.1093/jnci/djae072.

31. Li, H. & Durbin, R. Fast and accurate short read alignment with Burrows-Wheeler transform. Bioinformatics 25, 1754–1760 (2009).

32. Jones, D. et al. cgpCaVEManWrapper: Simple Execution of CaVEMan in Order to Detect Somatic Single Nucleotide Variants in NGS Data. Curr. Protoc. Bioinformatics 56, 15.10.1–15.10.18 (2016).

33. Ye, K., Schulz, M. H., Long, Q., Apweiler, R. & Ning, Z. Pindel: a pattern growth approach to detect break points of large deletions and medium sized insertions from paired-end short reads. Bioinformatics 25, 2865–2871 (2009).

34. Van Loo, P. et al. Allele-specific copy number analysis of tumors. Proc. Natl. Acad. Sci. U. S. A. 107, 16910–16915 (2010).

35. Nik-Zainal, S. et al. Landscape of somatic mutations in 560 breast cancer whole-genome sequences. Nature 534, 47–54 (2016).

36. Buels, R. et al. JBrowse: a dynamic web platform for genome visualization and analysis. Genome Biol. 17, (2016).

37. Dentro, S. C., Wedge, D. C. & Van Loo, P. Principles of Reconstructing the Subclonal Architecture of Cancers. Cold Spring Harb. Perspect. Med. 7, (2017).

38. Sherry, S. T. et al. dbSNP: the NCBI database of genetic variation. Nucleic Acids Res. 29, 308–311 (2001).

39. Karczewski, K. J. et al. The mutational constraint spectrum quantified from variation in 141,456 humans. Nature 581, 434–443 (2020).

40. Adzhubei, I., Jordan, D. M. & Sunyaev, S. R. Predicting Functional Effect of Human Missense Mutations Using PolyPhen-2. Curr. Protoc. Hum. Genet. 0 7, Unit7.20 (2013).

41. Ng, P. C. & Henikoff, S. SIFT: Predicting amino acid changes that affect protein function. Nucleic Acids Res. 31, 3812–3814 (2003).

42. Benjamini, Y. & Hochberg, Y. Controlling the False Discovery Rate: A Practical and Powerful Approach to Multiple Testing. J. R. Stat. Soc. Series B Stat. Methodol. 57, 289–300 (1995).

43. Coorens, T. H. H. et al. Lineage-Independent Tumors in Bilateral Neuroblastoma. N. Engl. J. Med. 383, 1860–1865 (2020).

44. Custers, L. et al. Somatic mutations and single-cell transcriptomes reveal the root of malignant rhabdoid tumours. Nat. Commun. 12, 1407 (2021).

45. Coorens, T. H. H. et al. Clonal origin of KMT2A wild-type lineage-switch leukemia following CAR-T cell and blinatumomab therapy. Nat Cancer 4, 1095–1101 (2023).

46. Moore, L. et al. The mutational landscape of human somatic and germline cells. Nature 597, 381–386 (2021).

47. Dobin, A. et al. STAR: ultrafast universal RNA-seq aligner. Bioinformatics 29, 15–21 (2013).

48. Anders, S., Pyl, P. T. & Huber, W. HTSeq—a Python framework to work with high-throughput sequencing data. Bioinformatics 31, 166–169 (2015).

49. Robinson, M. D., McCarthy, D. J. & Smyth, G. K. edgeR: a Bioconductor package for differential expression analysis of digital gene expression data. Bioinformatics 26, 139–140 (2010).

50. Aryee, M. J. et al. Minfi: a flexible and comprehensive Bioconductor package for the analysis of Infinium DNA methylation microarrays. Bioinformatics 30, 1363–1369 (2014).

